# Public Opinions towards COVID-19 in California and New York on Twitter

**DOI:** 10.1101/2020.07.12.20151936

**Authors:** Xueting Wang, Canruo Zou, Zidian Xie, Dongmei Li

## Abstract

**Background:** With the pandemic of COVID-19 and the release of related policies, discussions about the COVID-19 are widespread online. Social media becomes a reliable source for understanding public opinions toward this virus outbreak.

**Objective:** This study aims to explore public opinions toward COVID-19 on social media by comparing the differences in sentiment changes and discussed topics between California and New York in the United States.

**Methods:** A dataset with COVID-19-related Twitter posts was collected from March 5, 2020 to April 2, 2020 using Twitter streaming API. After removing any posts unrelated to COVID-19, as well as posts that contain promotion and commercial information, two individual datasets were created based on the geolocation tags with tweets, one containing tweets from California state and the other from New York state. Sentiment analysis was conducted to obtain the sentiment score for each COVID-19 tweet. Topic modeling was applied to identify top topics related to COVID-19.

**Results:** While the number of COVID-19 cases increased more rapidly in New York than in California in March 2020, the number of tweets posted has a similar trend over time in both states. COVID-19 tweets from California had more negative sentiment scores than New York. There were some fluctuations in sentiment scores in both states over time, which might correlate with the policy changes and the severity of COVID-19 pandemic. The topic modeling results showed that the popular topics in both California and New York states are similar, with “protective measures” as the most prevalent topic associated with COVID-19 in both states.

**Conclusions:** Twitter users from California had more negative sentiment scores towards COVID-19 than Twitter users from New York. The prevalent topics about COVID-19 discussed in both states were similar with some slight differences.

## Introduction

In December 2019, a novel and contagious coronavirus disease of 2019 (COVID-19) has been reported in China and continued spreading out massively. The cause of this pandemic is due to a novel coronavirus called SARS-CoV-2. According to the Centers for Disease Control and Prevention (CDC), the coronavirus could transmit between persons through close contact [1]. On January 30, 2020, COVID-19 became a Public Health Emergency of International Concern [2]. By June 22, 2020, the coronavirus has been confirmed in six continents with nearly nine million COVID-19 cases. The number of confirmed cases in the U.S. has been grown rapidly since early March. On March 7, New York declared a state of emergency after 89 confirmed cases, and since then, the confirmed cases continued increasing at an alarming rate. New York state was facing severe challenges in health care, including a shortage of protective gear and medical equipment, hospital overcrowding, and paramedic shortages. Meanwhile, California, as the most populous state in the U.S., was also facing the viral transmission, but less severe than what NY faced.

Discussions about the COVID-19 pandemic are widespread online, especially through social media, which offers powerful public platforms where people can share their opinions with large crowds. Twitter is an online microblogging and networking platform, and users can post and interact messages known as “tweets”. In past few years, Twitter data has been increasingly used for research. Previous study had used Twitter data to reveal insights from user-generated tweets about Ebola through textual analytics [3]. The outbreak of coronavirus in the U.S. has created an uproar in discussions about the pandemic on Twitter. A recent research collected a dataset of 9 million Twitter posts using the keywords “Coronavirus” and discussed the dynamics of coronavirus on Twitter [4]. However, little work has been reported on social media data analysis that focused on COVID-19 in the United States, since its outbreak on March 2020.

With the growth of the number of confirmed COVID-19 cases and the announcement of COVID-19 related policies, mentions of coronavirus-related topics and sentiments might vary over time. It’s important to understand how the perceptions of the pandemic related issues evolve over time, and whether they vary with different geolocations. Thus, in this study, we aimed to investigate the differences in sentiments and public opinions towards the outbreak of COVID-19 for twitter users between New York state and California state in the United States, and how these are related to the number of cases and policy changes. Our study provides valuable information about public attitude changes over time towards the emergence and outbreak of COVID-19, which could guide future research on exploring public concerns towards the pandemic.

## Methods

### Data Collection

We obtained COVID-19-related Twitter posts from March 5, 2020 to April 2, 2020, via Twitter streaming API. To generate only COVID-19-related posts, we used a list of keywords for Twitter streaming— “CORONA”, “corona”, “COVID19”, “covid19”, “covid”, “coronavirus”, “Coronavirus”, “CoronaVirus”, and “NCOV”. The dataset was then filtered by the same keyword set to remove any unrelated posts. Another filtering process was to remove the promotional and commercial posts. Since “Corona” is a beer brand in the market, it’s possible that the discussions in the posts about Corona were not related to the coronavirus. The keywords for filtering are “dealer”, “deal”, “supply”, “beer”, “drink”, “drank”, “drunk”, “store”, “promo”, “promotion”, etc. To identify tweets from California and New York, the last data filtering process is to separate tweets into two datasets — one dataset contains posts only from California state, another only from New York state, based on the geolocation information associated with each tweet. The keywords used to filter locations, such as “Los Angeles, CA”, “Monroe, New York”, are scraped from Wikipedia website (List of cities and towns) [5]. To prevent from missing locations while filtering, all the county and city names are considered into the keyword lists. By converting itemset into data frames and dropping the duplicates, we removed any duplicated items that share the same id number.

The number of confirmed COVID-19 cases in California and New York states each day from March 5, 2020 to April 2, 2020 were downloaded from the website of 1Point3Acres.com [6].

### Number of COVID-19 Tweets in Each State

In order to explore the trend of COVID-19-related tweets posted between March 5, 2020 and April 2, 2020, we calculated the number of tweets by days and hours for each state. To convert the date into the correct time zone, we parsed the date time into readable date format with the corresponding time zone. We calculated the average number of tweets posted by each hour within a 24-hour period.

### Sentiment Analysis

The VADER (Valence Aware Dictionary and sentiment Reasoner) was used for the sentiment analysis, which is a tool that is commonly used for sentiment analysis of social media data [7]. We applied the vaderSentiment package in python to calculate the sentiment score for each tweet and calculated the average sentiment scores by each day and each hour. For the text in each post, we extracted the compound score, which is normalized between −1 (negative extreme) and +1 (positive extreme). We calculated the average sentiment score within each hour in a 24-hour period for 29 days. In addition, a two-sample t-test was conducted to test whether there is a significant difference between average sentiment scores in California and New York based on the sentiment scores per minute.

### Topic Modeling Analysis

Topic Modeling is typically used to discover abstract “topics” in documents through statistical modeling [8]. To understand the most frequent topics that Twitter users were discussing about COVID-19, we applied Latent Dirichlet Allocation (LDA) for topic modeling analysis. To build the LDA model, several steps were conducted:

- Remove noises — emails, newline, extra spaces, distracting single quote and urls
- Tokenization — split texts into sentences and words, lower case the words and remove punctuation
- Create bigram and trigram models — convert two or three words frequently occurring together in the document (eg. difficulty_breathing, magic_johnson, catching_feelings)
- Remove stop words downloaded from nltk, and make bigrams and trigrams
- Lemmatization using spaCy — lemmatize the words to a normal form
- Create dictionary and corpus as LDA input

To identify the optimal number of topics, we calculated the coherence scores from 2 topics to 15 topics and considered the number of topics with the highest coherence score as the optimal number of topics. After building the LDA model, we applied pyLDAVis to visualize the information contained in the topic models. It’s a python built-in package that helps understand the meaning of each topic, the prevalence of each topic, and the correlation among topics. According to Sievert and Shirley’s study result, the optimal value of the weight parameter λ is 0.6 in the pyLDAVis method [9]. In our analysis, we adjusted the parameter to 0.6 and extracted the top ten frequent words in each topic. The summary of each topic was manually conducted and discussed among all coauthors.

## Results

### Temporal Analysis of COVID-19-related Tweets in California and New York

Figure 1 presents the number of newly reported COVID-19 cases each day from March 5, 2020 to April 2, 2020 in California and New York states. Since March 15, 2020, the number of confirmed cases started increasing at an alarming rate in New York, while the increasing rate in California is relatively low.

**Figure 1.**
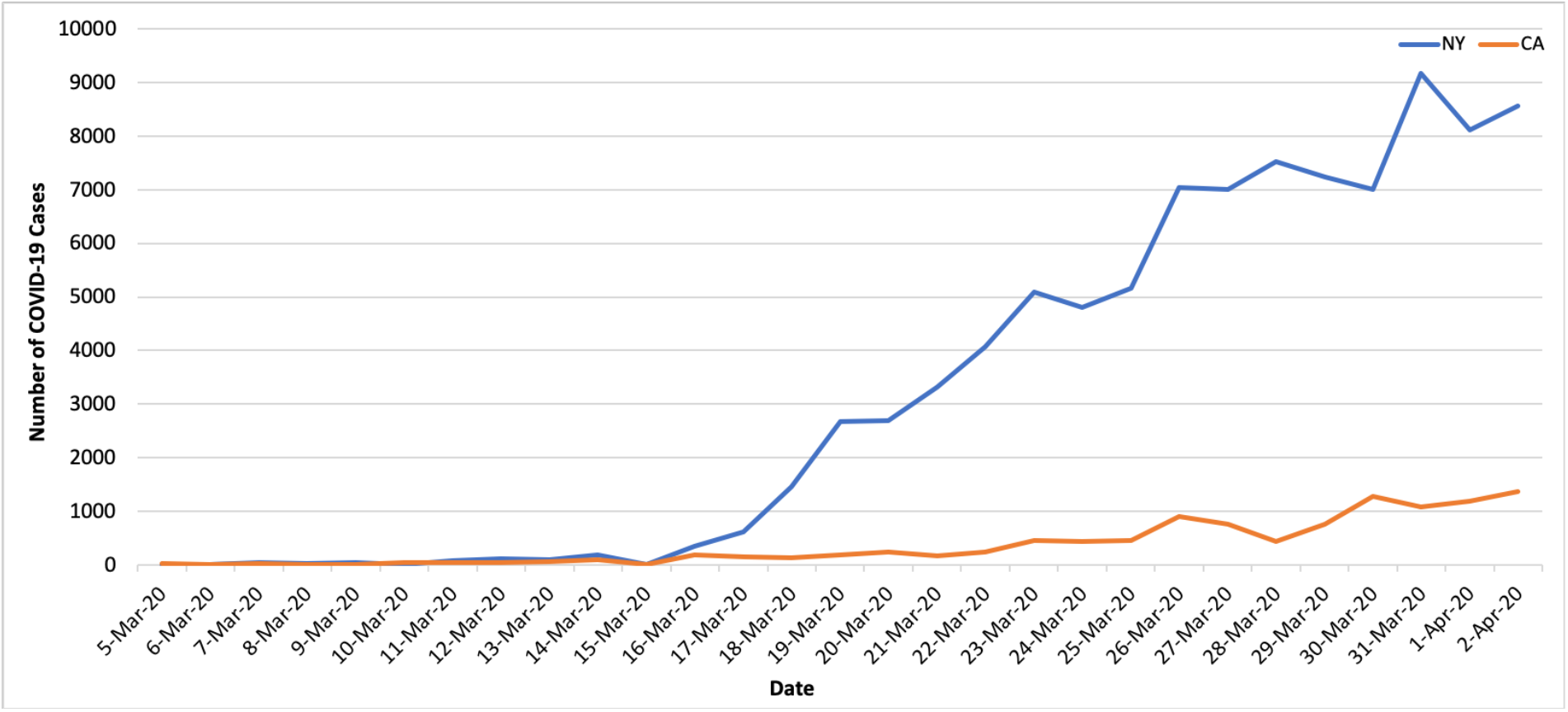
Number of confirmed COVID-19 cases in California and New York states.

To better analyze and compare the trend of number of COVID-19 tweets over time in California and New York from March 5, 2020 to April 2, 2020, we calculated the average number of COVID-19-related tweets each day for these two states. Overall, California Twitter users posted more COVID-19 tweets than New York Twitter users did (Figure 2). There were 2,336,593 tweets posted from California in total, while there were 1,385,745 tweets posted from New York. The overall trend in number of tweets in California and New York was very similar. Before March 11, 2020, the number of tweets increased every day in both states and started to drop until March 15, 2020. After that, the number of COVID-19 tweets remains relatively steady. In addition, in the 24-hour period, the average number of tweets began to rise from 4:00 AM in both states (Supplemental Figure 1). In New York, the peak time for the number of COVID-19 tweets is 8:00 PM. In California, the peak time is around 9:00 PM to 10:00 PM.

**Figure 2.**
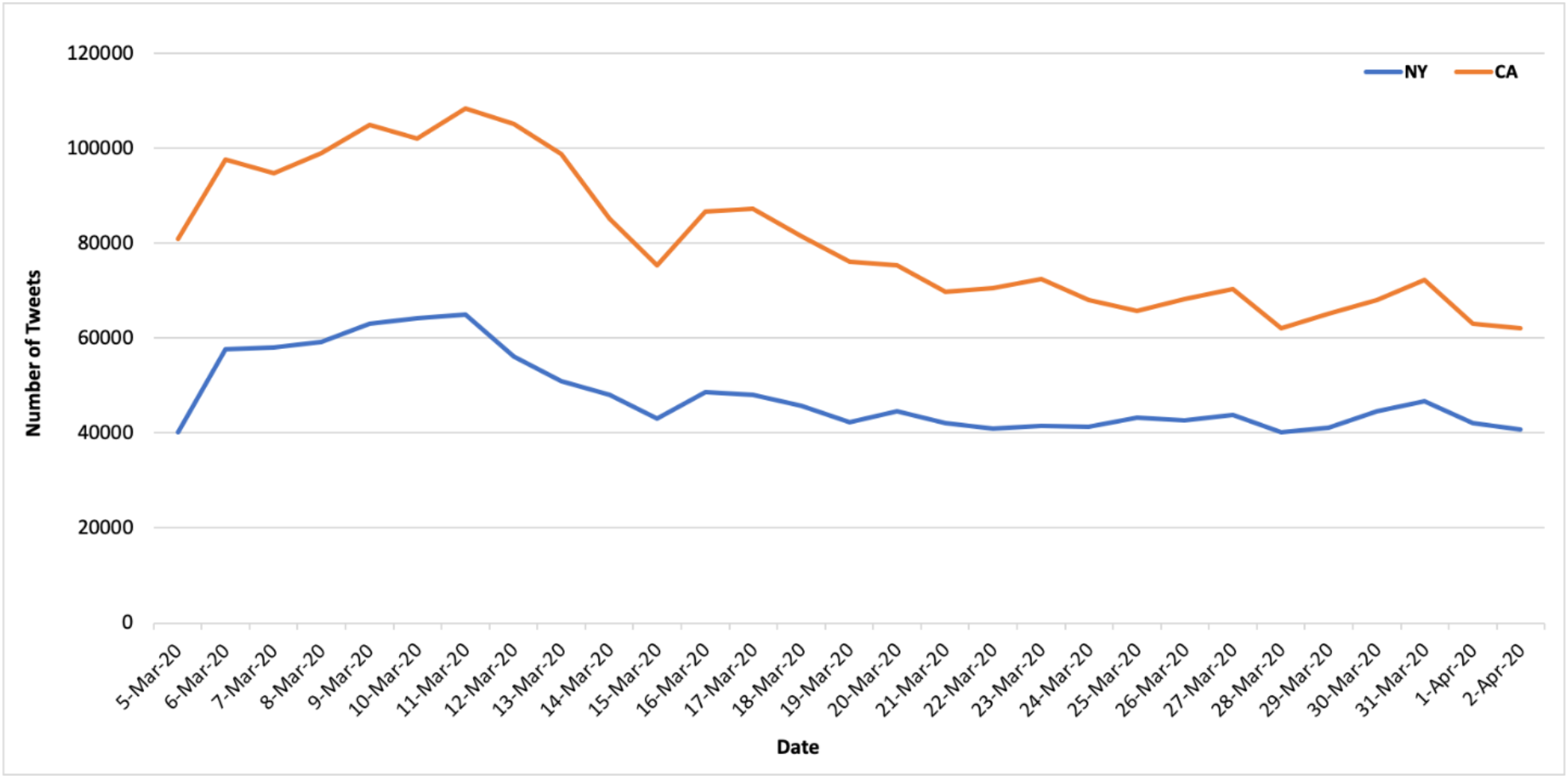
Number of COVID-19 tweets in California and New York over time.

### Different Sentiments towards COVID-19 between California and New York Twitter Users

A sentiment analysis was conducted to understand public attitudes toward the number of COVID-19 cases and the policy changes. We calculated average sentiment scores for COVID-19 tweets from California and New York between March 5, 2020 and April 2, 2020. Overall, both states had negative sentiment scores towards COVID-19. The average sentiment score for California was −0.042 (Standard Error = 0.0004), while for New York, it was −0.033 (Standard Error = 0.0003). A two-sample *t-*test showed that the difference in average sentiment scores between California and New York was significant (p-value < 0.001).

The daily average sentiment scores in both New York and California varied over time (Figure 3). The lowest sentiment score (−0.093) occurred on March 7, 2020 when New York announced the state of emergency (Figure 3A). The highest sentiment score (0.003) happened on March 25, 2020 when the Congress agreed on a $2 trillion virus relief package bill. On March 29, when New York surpassed 1000 death toll, the sentiment score was −0.058. On the other hand, average sentiment score in California has a similar trend as in New York (Figure 3B). For example, on March 22, 2020 the White House gave California coronavirus crisis disaster designation, and the sentiment score was down to −0.058.

**Figure 3.**
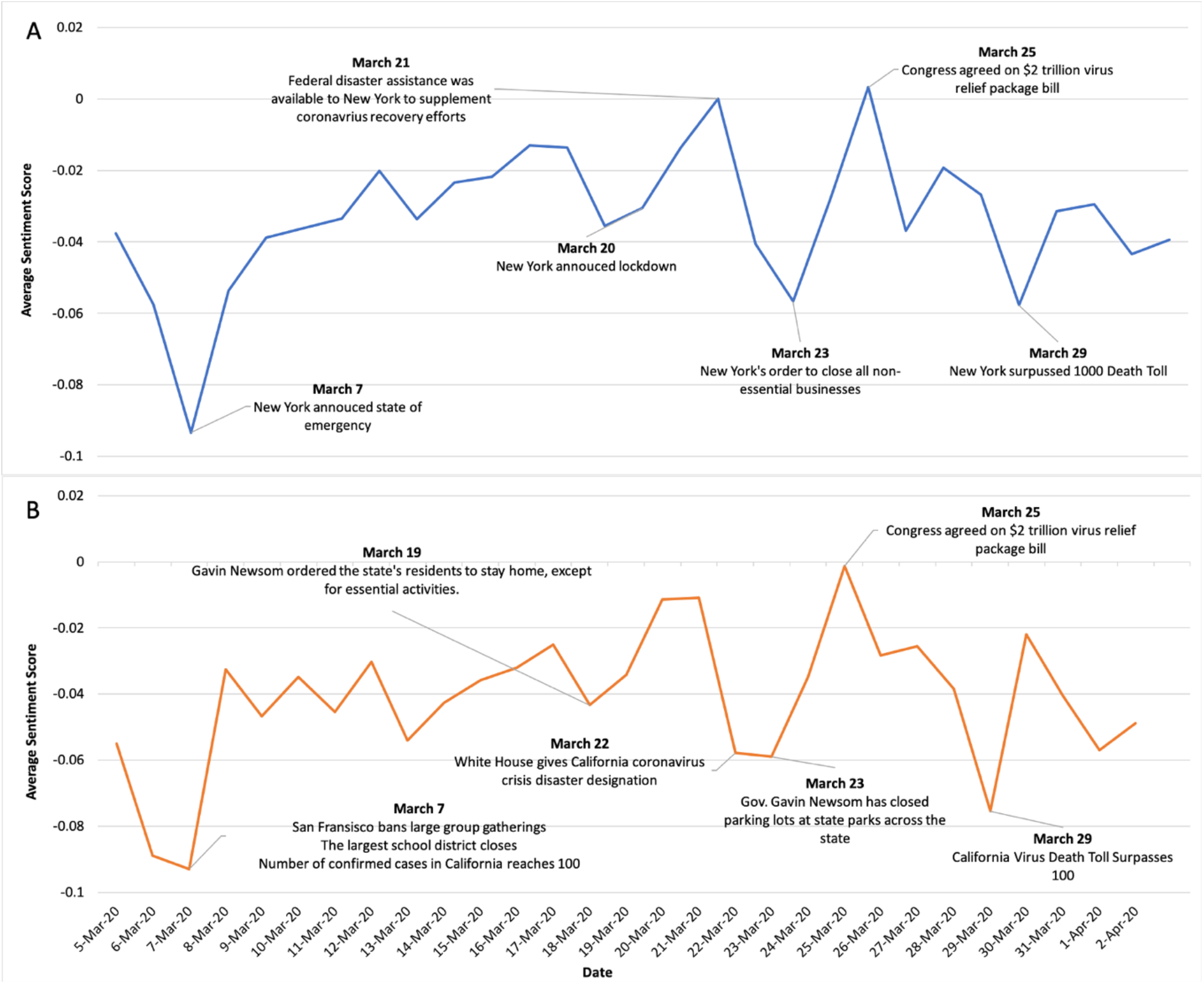
Average sentiments scores towards COVID-19 over time. (A) New York; (B) California

In the 24-hour period, New York showed a clear peak on the hourly average sentiment score, with the highest sentiment score of −0.018 at 11:00AM (Supplemental Figure 2). In contrast, California did not show a clear peak. In addition, twitter users from both states showed more negative sentiments towards COVID-19 from midnight to early morning.

### Topics about COVID-19 Tweets in New York and California

The LDA topic modeling was applied to understand the general topics in COVID-19 tweets. Based on the coherence scores, we found that with New York data the optimal number of topics was 13 while with California data it was 14. We then summarized each topic with top 10 frequent words (Table 1).

**Table 1.** Topics associated with COVID-19 tweets from New York and California.

| New York |  |  | California |  |  |
| --- | --- | --- | --- | --- | --- |
| Topics | %Token | Keyword | Topics | %Token | Keyword |
| Protective Measures | 7.73 | work, stop, home, amp, stay, spread, hand, mask, face, people | Protective Measures | 7.20 | work, stop, home, stay, sick, hand, leave, mask, pay, face |
| Pandemic's impact | 7.72 | sick, leave, news, pay, travel, emergency, global, business, impact, worker | State orders | 7.18 | break, health, state, public, testing, order, announce, emergency, official, coronavirus |
| Government and Policy | 7.71 | trump, health, care, public, official, break, crisis, hold, company, president | Public concern | 7.16 | people, kill, fact, risk, infect, amp, hit, lot, avoid, run |
| Medical support | 7.70 | covid, doctor, fight, put, pandemic, medical, risk, expert, nurse, send | Bill reliefs | 7.16 | family, school, pass, vote, plan, relief, business, provide, student, bill |
| Public concern | 7.70 | people, due, start, cancel, hear, kill, medium, fear, concern, infect | Government and Policy | 7.15 | trump, response, medium, lie, crisis, president, handle, early, claim, release |
| Government response | 7.69 | government, response, outbreak, order, story, follow, threat, release, lead, part | Hospital situation | 7.14 | die, day, year, hospital, patient, ago, hour, week, flu, night |
| Impact on life | 7.68 | today, family, job, plan, friend, lose, hour, pass, vote, night | Test results | 7.14 | test, positive, covid, feel, symptom, shit, life, find, follow, kit |
| School closure | 7.68 | week, call, close, country, school, month, chinese, ago, lie, world | Quarantine | 7.13 | today, care, show, live, quarantine, lose, read, share, covid, open |
| Hospital situation | 7.68 | patient, hospital, talk, live, watch, end, share, question, video, show | Confirmed cases and deaths | 7.13 | case, death, report, number, confirm, country, world, update, late, high |
| Testing and vaccine | 7.68 | test, positive, day, testing, find, feel, free, symptom, covid, vaccine | Vaccine and Immunization | 7.13 | week, travel, vaccine, immune, system, point, kid, problem, expose, thousand |
| The impact of COVID-19 | 7.67 | virus, corona, die, year, quarantine, flu, man, play, woman, kid | Impact on public resources | 7.12 | due, cancel, close, concern, fear, outbreak, real, play, shut, food |
| Confirmed cases and deaths | 7.67 | case, death, state, report, confirm, number, update, late, total, high | Global news | 7.11 | call, fight, bad, chinese, month, news, pandemic, government, question, expert |
|  |  |  | The pandemic of COVID-19 | 7.12 | virus, corona, spread, put, back, continue, give, money, start, catch |

As shown in Table 1, topics discussed in COVID-19 tweets in both New York and California were very similar, among which the most prevalent topic was “Protective measures”. In addition, concerns about the COVID-19 pandemic and government actions, such as “Public concerns” and “Government and policy”, were other popular topics in both states.

## Discussion

In this study, we showed that from March 5 to April 9 in 2020, the number of COVID-19 tweets in California was almost twice of that in New York. California had a significantly lower sentiment score towards COVID-19 than New York. In addition, we showed that the sentiment scores towards COVID-19 in both California and New York varied over time, and the policy announcements and number of confirmed cases might be the major drives for these sentiment changes. Twitter users from both states discussed similar topics, with the “protective measures” as the most popular topic, followed by “Pandemic’s impact” and “Government policy”.

While we showed that the sentiment scores varied over time in both California and New York, the highest and lowest sentiment scores correlated with some important policy announcements and the severity of COVID-19. On March 7, Governor Cuomo declared a State of Emergency in New York, and the sentiment score on that day was the lowest (−0.093). One possible explanation is that people expressed serious concerns about the COVID-19 pandemic. The word “emergency”, along with “sick”, “leave”, “business”, “worker”, suggests that people were worried about their businesses and not being able to work. Meanwhile, on the same day, the average sentiment score in California was the lowest, which coincide with the policies including the largest school district was closed, and San Francisco banned large group gatherings. Another interesting observation is that on March 25, 2020, the average sentiment scores in both states reached the highest point. The major event happened on that day was that a $2 trillion coronavirus relief package was passed to address the economic impacts caused by the pandemic. It can be inferred that since the pandemic has forced a lot of people losing their jobs and not being able to maintain their livings, the announcement of relief bill could relieve some financial stresses, which led to less negative sentiment scores in both states. In addition, we found that the number of confirmed cases and deaths might correlate with the public’s sentiment. For instance, on March 7, the number of confirmed cases in California reached 100 [10]; on March 29, New York surpassed 1000 death tolls [11], and California surpassed 100 death tolls [12]. These numbers might be the drives for more negative sentiment scores in both states.

Since New York had much more confirmed cases and deaths than California in March 2020, we would expect that the average sentiment score in New York would be lower than in California. However, our results showed that California had a lower sentiment score (−0.042) than New York (−0.033), and the difference is significant. One explanation for this is that on March 21, 2020, New York began gathering ventilators across the state. The federal disaster assistance might bring people who were in pessimistic situations some hopes. A recent research by WalletHub about the unemployment rate showed that New York had a 1569.69% increase in the unemployment claims since the start of the COVID-19 pandemic, and California had a 1143.54% increase, much less than New York [13]. Thus, the $2 trillion relief bill happened on March 25 could provide people in New York financial support more significantly. Another reasonable speculation could be that as the most populous state in the U.S., California people might be more worried about the potential high infection rate.

Our result on topic modeling analysis showed that there was no major difference between the topics discussed in California and New York. However, the popularity of each topic was different between two states. The topic that had the highest token percentage in both states was related to protective measures. They shared the same keywords such as “work”, “stop”, “home”, “stay”, “hand”, “mask”. It can be inferred that these tweets were talking about staying home, washing hands, and wearing masks. The second prevalent topic in New York was about the impact that pandemic brought, including keywords such as “business”, “travel”, “worker”. Since New York had far more confirmed cases and deaths, the negative impacts were more severe, and people might discuss more about the harm to businesses and traveling restrictions. In contrast, tweets from California talked less about working impacts according to the topic modeling keywords, presumably due to less serious situations. Topics related to government and policy seem to be the next mostly discussed categories. Keywords including “trump”, “president”, and “crisis” appeared in tweets from both states with high frequencies. Other than the prevention and impacts of COVID-19, people paid significant attention to the government policy and president’s actions, which might partially explain the fluctuation of sentiment scores. Hospital environment and medical support were other popular topics in both states.

Although social media provides first-hand users’ perspectives, it does not provide controlled variables. We don’t have the user demographic information, including age, gender, and education, etc. Twitter users can’t represent the whole population, since about only 20% of US adults are currently using Twitter. Furthermore, our study only focused on the beginning of COVID-19 epidemic in the US, and the story might evolve later.

This study used social media data from Twitter to analyze the sentiment scores and public opinions towards the emergence of COVID-19 in California and New York states in the United States at the beginning of the COVID-19 epidemic. This study showed how COVID-19 affected people’s attitudes from different states in a timely manner. The results from this study could provide guidelines for future social media research on the associations between the outbreak of COVID-19 and public opinions, as well as valuable suggestions on future pandemic research.

## Data Availability

We obtained COVID-19-related Twitter posts from March 5, 2020 to April 2, 2020, via Twitter streaming API. Twitter data are available upon request.

## Acknowledgements

The project described in this publication was supported by the University of Rochester Clinical and Translational Science Award UL1 TR002001 from the National Center for Advancing Translational Sciences of the National Institutes of Health (DL).

---

**Supplemental Figure 1.**
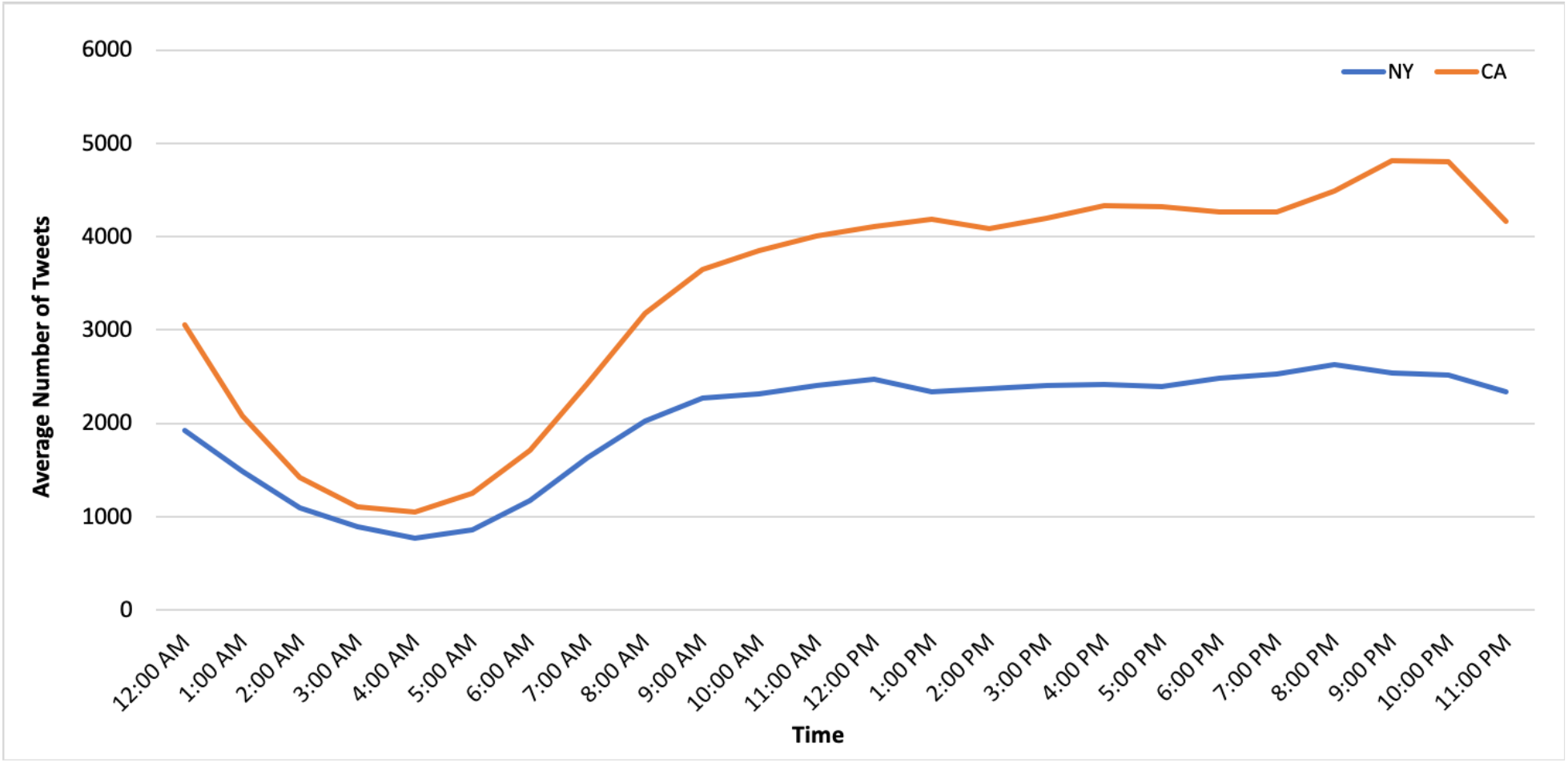
Number of COVID-19 tweets within a 24-hour period in California and New York.

**Supplemental Figure 2.**
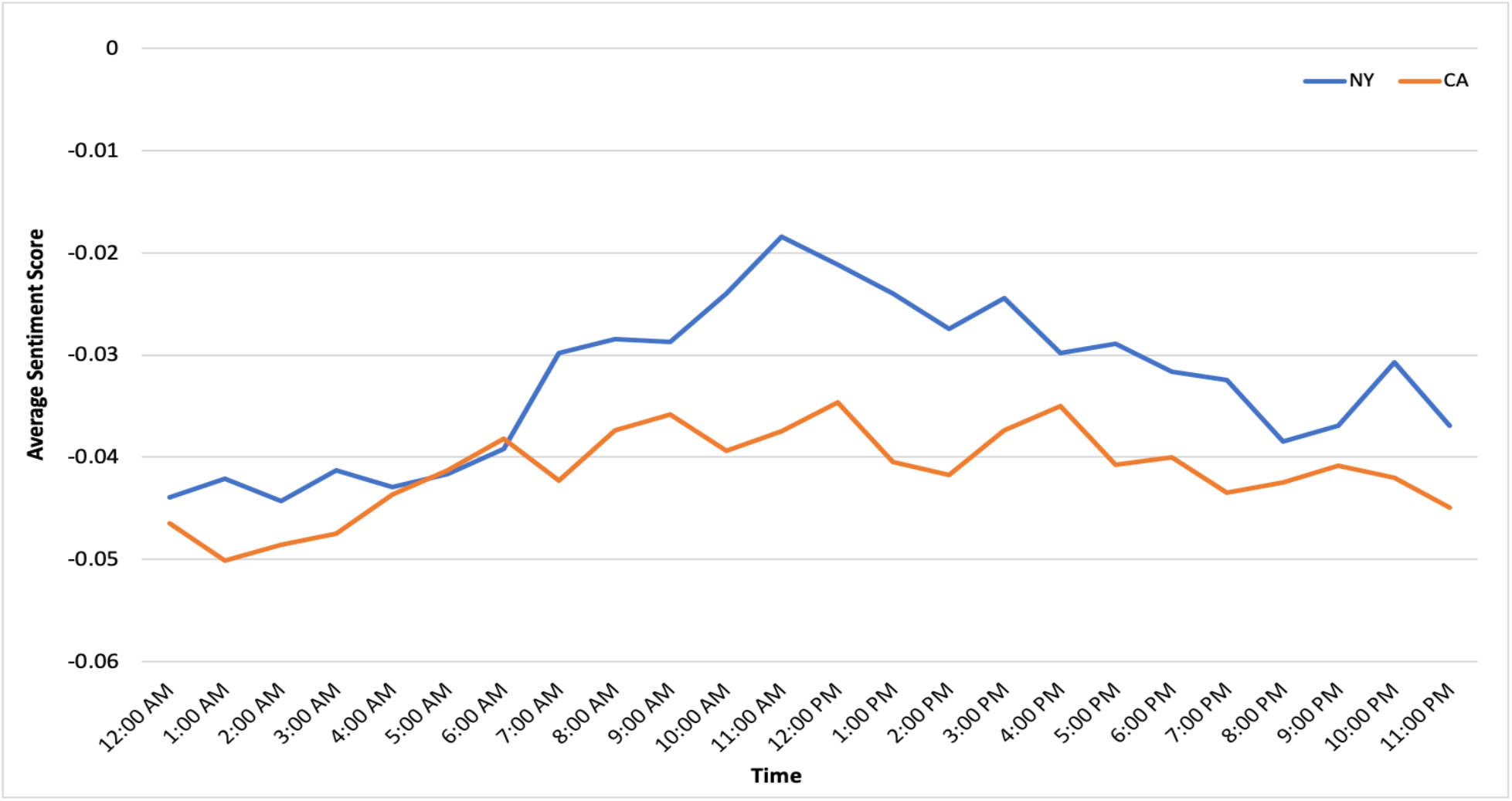
Sentiment scores towards COVID-19 in 24-hour period in California and New York.

## References

[1] CDC. “Coronavirus (COVID-19) Frequently Asked Questions.” Centers for Disease Control and Prevention, Centers for Disease Control and Prevention, 2 June 2020, www.cdc.gov/coronavirus/2019-ncov/faq.html.

[2] World Health Organization. “Coronavirus Disease (COVID-19) – Events as They Happen.” World Health Organization, World Health Organization, 2020, www.who.int/emergencies/diseases/novel-coronavirus-2019/events-as-they-happen.

[3] Lazard, Allison J., et al. “Detecting Themes of Public Concern: A Text Mining Analysis of the Centers for Disease Control and Prevention’s Ebola Live Twitter Chat.” American Journal of Infection Control, vol. 43, no. 10, 2015, pp. 1109–1111., doi:10.1016/j.ajic.2015.05.025.

[4] Aguilar-Gallegos, Norman, et al. “Dataset on Dynamics of Coronavirus on Twitter.” Data in Brief, vol. 30, 2020, p. 105684., doi:10.1016/j.dib.2020.105684.

[5] “List of Cities and Towns in California.” Wikipedia, Wikimedia Foundation, 8 May 2020, en.wikipedia.org/wiki/List_of_cities_and_towns_in_California.

[6] “Global COVID-19 Tracker & Interactive Charts: Real Time Updates & Digestable Information for Everyone.” 1Point3Acres, coronavirus.1point3acres.com/.

[7] Hutto, C.J. & Gilbert, E.E. (2014). VADER: A Parsimonious Rule-based Model for Sentiment Analysis of Social Media Text. Eighth International Conference on Weblogs and Social Media (ICWSM-14). Ann Arbor, MI, June 2014.

[8] Li, Susan. “Topic Modeling and Latent Dirichlet Allocation (LDA) in Python.” Medium, Towards Data Science, 1 June 2018, towardsdatascience.com/topic-modeling-and-latent-dirichlet-allocation-in-python-9bf156893c24.

[9] Sievert, Carson, and Kenneth Shirley. “LDAvis: A Method for Visualizing and Interpreting Topics.” Proceedings of the Workshop on Interactive Language Learning, Visualization, and Interfaces, 2014, doi:10.3115/v1/w14-3110.

[10] The New York Times. “California Coronavirus Map and Case Count.” The New York Times, The New York Times, 1 Apr. 2020, www.nytimes.com/interactive/2020/us/california-coronavirus-cases.html.

[11] MassLive, The Associated Press |. “Coronavirus Death Toll in New York Surpasses 1,000, Less than 3 Weeks after 1st Reported Death.” Masslive, 30 Mar. 2020, www.masslive.com/coronavirus/2020/03/coronavirus-death-toll-new-york-surpasses-1000-covid-19-deaths.html.

[12] Eby, Kate. “Coronavirus Timeline: Tracking Major Moments of COVID-19 Pandemic in San Francisco Bay Area.” ABC7 San Francisco, 16 June 2020, abc7news.com/timeline-of-coronavirus-us-coronvirus-bay-area-sf/6047519/.

[13] McCann, Adam. States Hit Most by Unemployment Claims. WalletHub 2020. Print.

